# Inequalities in Colorectal Cancer Screening: Combining MAIHDA with Difference-in-Differences to Assess Programme Effects Across Population Subgroups

**DOI:** 10.64898/2026.07.16.26358242

**Authors:** Vladimir Jolidon, Katrijn Delaruelle, Ichiro Kawachi, Stéphane Cullati, Andrew Bell, Daniel Holman

**Affiliations:** Department of Social and Behavioral Sciences, Harvard T.H. Chan School of Public Health, Boston, MA, USA; School of Sociological Studies, Politics and International Relations, University of Sheffield, 2 Whitham Road, Sheffield, S10 2AH, United Kingdom; Population Health Laboratory (#PopHealthLab), University of Fribourg, Route des Arsenaux 41, 1700 Fribourg, Switzerland; Department of Sociology, Ghent University, Sint-Pietersnieuwstraat 41, 9000 Ghent, Belgium; Institute of Sociological Research, University of Geneva, 1205 Geneva, Switzerland; Swiss Centre of Expertise in Life Course Research LIVES, Lausanne-Geneva, Switzerland; Sheffield Methods Institute, School of Education, University of Sheffield, 2 Whitham Road, Sheffield, S10 2AH, United Kingdom

**Keywords:** Fecal Occult Blood Test, Colorectal Neoplasms, Diagnostic Screening Programs, Healthcare Disparities, Multilevel Analysis, Sociodemographic Factors, Quasi-Experimental Studies

## Abstract

**Background:** Research consistently shows that colorectal cancer (CRC) screening uptake is socially patterned; however, sociodemographic determinants are usually analysed separately, overlooking how multiple social conditions jointly shape inequalities. This also applies to policy research, where heterogeneity in screening programme effects remains underexplored.

**Methods:** Using data from the European Health Interview Survey (2014 and 2019; n=201,214; 24 countries), we applied Multilevel Analysis of Individual Heterogeneity and Discriminatory Accuracy (MAIHDA) to analyse CRC screening uptake across 72 subgroups defined by sex, education, living arrangement and employment. To assess heterogeneity in screening programme effects, we combined MAIHDA with difference-in-differences (MAIHDA-DiD).

**Results:** MAIHDA revealed inequalities in uptake: lower- and middle-educated men, whether employed or unemployed, had the lowest uptake, whereas men and women not living alone, retired or living with disability, had the highest uptake. Lower-educated homemaker women were the only female group with below-average uptake. MAIHDA-DiD showed that programmes increased overall uptake but did not produce larger gains among groups with lower pre-intervention uptake, and therefore did not reduce inequalities. Instead, programmes generated above-average increases among groups with higher pre-intervention uptake, particularly lower- and middle-educated men and women not living alone and retired. Living arrangement explained more variation in programme effects than other factors, with individuals living alone benefiting less from the programmes.

**Conclusion:** CRC programmes did not reduce (and may have widened) inequalities, underscoring the need for equity-focused strategies in population-based screening. By extending MAIHDA with difference-in-differences, this study introduces a novel approach for evaluating heterogeneous policy effects in public health.

## Introduction

Colorectal cancer (CRC) is among the most diagnosed cancers and a leading cause of cancer-related mortality in Europe (1, 2). Evidence-based guidelines recommend systematic screening for individuals aged 50-74, and organised screening has contributed to reductions in CRC incidence and mortality through early detection and treatment (3, 4). The Council of the European Union recommends organised CRC screening programmes with systematic invitations, preferably using a faecal immunochemical test (FIT) (5). However, programme implementation remains uneven, and some countries still rely on opportunistic screening, offered during healthcare visits or initiated by individuals, without centralised invitation systems (6, 7).

Assessing social inequalities in screening uptake is important to ensure all population groups benefit equally from CRC prevention. Numerous studies document clear social patterning; for instance, individuals with lower educational attainment consistently underuse cancer screening (8, 9). Research has also shown that unemployed individuals tend to participate less in screening, while retirees participate more, although findings are mixed and some studies report no effect of employment status (10). Because socioeconomic position is multidimensional, the effect of employment status likely overlaps with other sociodemographic characteristics, suggesting that it should be examined in combination with other factors rather than in isolation. Social support has also been highlighted as a key determinant: a European study showed that living alone negatively affected CRC screening uptake, particularly among men (11). However, most research has examined determinants separately, overlooking how they jointly shape inequalities. This single “risk-factor” approach, common in health inequalities research, isolates individual variables with limited attention to their interactions (12), thereby concealing how multiple factors shape social positions, cumulative (dis)advantages and subgroup-specific inequalities.

A novel approach has gained recognition for studying inequalities at the intersection of multiple social factors: Multilevel Analysis of Individual Heterogeneity and Discriminatory Accuracy (MAIHDA) (13). MAIHDA nests individuals within “strata” defined by combinations of sociodemographic characteristics (e.g. gender, age, ethnicity), reflecting that individuals within the same stratum share similar social contexts, such as access to resources or exposure to discrimination. These characteristics are not only individual attributes but markers of broader social structures that shape opportunities and constraints (14). Commonly considered factors include gender, ethnicity, socioeconomic position and age, as they represent major axes of social inequality and social identity (15). MAIHDA has been applied to outcomes such as body mass index (13), depression (16) and alcohol consumption (17). However, to our knowledge, MAIHDA has not yet been applied to preventive healthcare utilisation such as CRC screening, despite evidence of unequal uptake.

In addition to social factors, cancer screening programmes are a key determinant of inequalities in uptake. Comparative studies suggest that countries with organised programmes have smaller socioeconomic inequalities in uptake than those relying on opportunistic screening (18-20). However, these studies did not use causal designs, and observed differences could reflect pre-existing inequalities or other contextual factors. Several European countries have introduced CRC programmes with mailed FIT kits, enabling home-based testing and postal return. While such programmes have consistently increased overall screening uptake (21), their effects on inequalities in uptake across intersecting sociodemographic groups remain understudied.

Here, we assess inequalities in CRC screening uptake across population subgroups defined by multiple intersecting social conditions. Departing from prior research that has analysed determinants in isolation, we apply MAIHDA to capture their interplay. Exploiting a quasi-experimental context in which four European countries implemented nationwide CRC programmes between 2014 and 2019, we combine MAIHDA with difference-in-differences methods to evaluate programme effects on screening uptake and inequalities.

## Methods

We used population-based data from the European Health Interview Survey (EHIS), wave 2 (EHIS-2, 2013-2015) and wave 3 (EHIS-3, 2018-2020). EHIS is a nationally administered cross-sectional survey of individuals aged 15 and older living in private households.

The study included 24 countries with data available in both EHIS-2 and EHIS-3, providing the two time points required for the difference-in-differences design (Supplementary Material Table S.1). The analytical sample was restricted to individuals aged 50-74, in line with CRC screening recommendations (6) and the target population of national programmes. In Belgium, Luxembourg, and the Netherlands, where programmes targeted ages 55-74, samples were restricted accordingly. The final sample included 201,214 respondents after excluding those with missing data on the outcome (n=6,772, 3.22%) or strata-defining variables (n=2,778, 1.32%).

## Measures

### Outcome

EHIS asked respondents when they last had a faecal occult blood test (within the past 12 months, 1 to less than 2 years ago, 2 to less than 3 years ago, more than 3 years ago, never). Our outcome was uptake within the past two years, versus screening more than two years ago or never screened, in line with European guidelines recommending biennial stool-based screening (6).

### Social strata

We defined 72 strata by combining sex (male, female), education (lower, middle, higher), living arrangement (living alone, not alone) and employment status (employed, unemployed, homemaker, disability, retired, other inactive). Sex was measured using the EHIS variable capturing sex assigned at birth (gender identity was not measured). In EHIS, education followed the 2011 International Standard Classification of Education (ISCED-2011). We recoded educational attainment into ‘lower’ (ISCED 0-2, primary or lower secondary education), ‘middle’ (ISCED 3-4, upper secondary or post-secondary non-tertiary education) and ‘higher’ (ISCED 5-8, tertiary education). We used employment status as provided; except that ‘other inactive person’ and ‘student’ were combined into ‘other inactive’ due to their sample sizes. Although MAIHDA studies often include age as a stratifying variable, retirement status in our analysis captured key age-related differences and an important life-course transition relevant to CRC screening. Therefore, age was not included as a stratifying variable.

### The intervention: CRC screening programme with mailed FIT kit

Between EHIS-2 and EHIS-3, Belgium, Denmark, Luxembourg and the Netherlands implemented nationwide CRC programmes that mailed FIT kits to all individuals in the target age group. We classified respondents from these countries (n=16,643) as the intervention group, while respondents from the remaining 20 European countries (n=184,571) without such programmes during the study period formed the comparison group (Supplementary Material Table S.1). This classification was based on published literature and policy documentation (6, 7, 22, 23).

### Statistical analysis

First, we performed descriptive analyses of the analytical sample. Second, we applied MAIHDA to estimate CRC screening uptake across social strata in the pre-intervention period (EHIS-2). Model 1a was specified as:

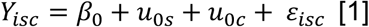

where *Yisc* denotes screening uptake for individual *i* in stratum *s* and country *c*, *β0* represents the estimated population mean, specifically the “precision-weighted grand mean” (PWGM), *u0s* the random intercept measuring the stratum-level deviations from the mean, and *u0c* is a country-level random intercept accounting for clustering. Within the MAIHDA framework, between-stratum heterogeneity can be decomposed into additive and multiplicative effects of the strata (15). To assess this, we re-estimated Model 1a including all stratum-defining variables as main effects (Model 1b).

Next, we combined MAIHDA with difference-in-differences to assess programme effects across strata. Model 2a included variables for programme (0 = no programme; 1 = programme), time (0 = pre-intervention; 1 = post-intervention) and their interaction (DiD). To account for unobserved heterogeneity across countries and survey waves, we included a country-time random intercept. This specification captures dependence across countries and time, helping to prevent downward bias in standard errors while isolating the programme effect from other concurrent influences (24). The equation is as follows:

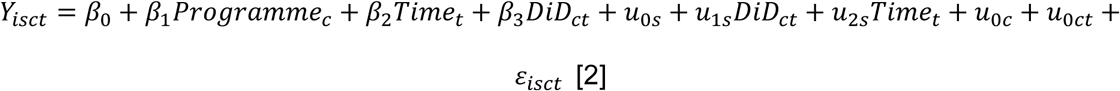

Here, *β*_1_ captures pre-intervention differences in screening uptake between programme and comparison countries, while *β*_2_ reflects temporal changes for the comparison group. The key parameters of interest are the DiD coefficient *β*_3_, estimating the average programme effect, and the random slopes *u1s* for the DiD term, capturing heterogeneity in programme effects across strata. Including random slopes for time improved model fit, whereas programme random slopes worsened model performance and had negligible variance. To avoid over-parameterisation, only time random slopes were retained in the model. In all models, random coefficients were assumed to be normally distributed.

For Model 1a, we derived predicted stratum-level deviations from the overall mean pre-intervention uptake (random intercepts), and for Model 2a, predicted stratum-level deviations from the overall programme effect (random slopes for the DiD term). These were reported together in a single table.

To examine which sociodemographic factors explained most of the variation in programme effects across strata, we estimated additional models interacting the DiD term with each stratum-defining variable separately. We calculated the Proportional Change in Variance (PCV) of the DiD random slope to quantify the extent to which each factor accounted for between-stratum heterogeneity in programme effects.

Models were estimated using linear probability models with an identity link function, an approach commonly applied in difference-in-differences analyses, as the linear DiD estimator is consistent for the average treatment effect even with a binary outcome (25). Linear probability models are often preferred because they preserve the DiD additive structure, providing directly interpretable effect sizes as percentage-point changes. In contrast, in logit models, the interaction term reflects effects on the latent index rather than on the outcome probability and therefore does not represent the actual difference-in-differences in outcome probabilities (26).

Difference-in-differences assumptions were assessed using pre-trend, placebo and falsification tests (Appendix A in Supplementary Material). Overall, these analyses support the validity of the DiD identification strategy. Due to limited pre-intervention data, the parallel trends assumption could be formally tested only for Belgium, where it was supported.

Nevertheless, similar pre-intervention screening uptake levels between intervention and comparison groups support the plausibility of parallel trends in other countries, and the falsification and placebo tests further corroborate the identification strategy, suggesting that the estimated programme effects are unlikely to be driven by broader shifts in healthcare use or other time-varying confounders.

Analyses were performed using StataSE 18.

## Results

### Sample characteristics

Table 1 summarises the analytical sample. 26.2% of respondents reported stool-based CRC screening in the past two years. Women represented 54.2% of the sample, the largest educational category was upper secondary (42.0%), and the largest occupational groups were employed (41.2%) and retired (41.8%). In EHIS-2 and EHIS-3, strata sizes ranged from 9 to 16,687 respondents (Supplementary Table S.2), with only four strata (1.3%) containing fewer than 50 respondents.

**Table 1.** Characteristics of analytical sample, EHIS-2 and EHIS-3 (N = 201,214)

|  | N | % |
| --- | --- | --- |
| **Stool-based screening in the past 2 years** |  |  |
| More than 2 years ago | 148,494 | 73.8 |
| In the past 2 years | 52,720 | 26.2 |
| **Sex** |  |  |
| Female | 109,161 | 54.2 |
| Male | 92,053 | 45.8 |
| **Education level** |  |  |
| Lower | 70,319 | 35.0 |
| Middle | 84,571 | 42.0 |
| Higher | 46,324 | 23.0 |
| **Living arrangement** |  |  |
| Living alone | 43,657 | 21.7 |
| Not living alone | 157,557 | 78.3 |
| **Employment status** |  |  |
| Employed | 82,870 | 41.2 |
| Unemployed | 8,998 | 4.5 |
| Retired | 84,027 | 41.8 |
| Disability | 7,035 | 3.5 |
| Homemaker | 13,932 | 6.9 |
| Other inactive | 4,352 | 2.2 |

EHIS = European Health Interview Survey

### Pre-intervention screening uptake across subgroups

MAIHDA regression results for the pre-intervention period are reported in Table 2. In Model 1a, the intercept corresponds to an overall predicted screening uptake of 18%. In Model 1b, fixed effect estimates show the additive contributions of the stratifying variables. Male sex, lower education, living alone, and being employed, unemployed, a homemaker or other inactive were associated with lower uptake, whereas disability was associated with higher uptake. In this fully adjusted model, the stratum-level variance reduced to near zero, indicating that almost all between-stratum differences were explained by additive rather than multiplicative effects. Only two strata (lower- and middle-educated retired men not living alone) showed statistically significant, though small (about one percentage point), positive residual multiplicative effects.

**Table 2.** MAIHDA regression estimates for past-two-year screening uptake, EHIS-2 (N = 93,571)

|  | Model 1a |  | Model 1b |  |
| --- | --- | --- | --- | --- |
|  | Estimate | 95% CI | Estimate | 95% CI |
| **Fixed-effects** |  |  |  |  |
| Intercept | 0.1801 | 0.1273, 0.2328 | 0.2219 | 0.1687, 0.2751 |
| *Sex (ref.: female)* |  |  |  |  |
| male | - |  | -0.0080 | -0.0156, -0.0003 |
| *Education (ref.: high)* |  |  |  |  |
| low | - |  | -0.0231 | -0.0332, -0.0130 |
| middle | - |  | -0.0086 | -0.0183, 0.0012 |
| *Living arrangement (ref.: not alone)* |  |  |  |  |
| alone | - |  | -0.0193 | -0.0273, -0.0113 |
| *Employment status (ref.: retired)* |  |  |  |  |
| employed | - |  | -0.0299 | -0.0386, -0.0211 |
| unemployed | - |  | -0.0497 | -0.0634, -0.0360 |
| disability | - |  | 0.0203 | 0.0055, 0.0351 |
| homemaker | - |  | -0.0368 | -0.0516, -0.0221 |
| other inactive | - |  | -0.0397 | -0.0589, -0.0204 |
| **Random intercept variances** |  |  |  |  |
| Country level | 0.0170 |  | 0.0170 |  |
| Stratum level | 0.0007 |  | 0.0001 |  |
| Individual level residual variance | 0.1455 |  | 0.1455 |  |

Note: Multilevel models with individuals cross-classified by sociodemographic strata (n = 72) and countries (n = 24), i.e. random intercepts for both strata and countries.
MAIHDA = Multilevel Analysis of Individual Heterogeneity and Discriminatory Accuracy; DiD = Difference-in-Differences

Table 3 presents the predicted stratum-level deviations from the overall mean (random intercept residuals), showing the 15 strata with the highest and lowest predicted uptake (full results in Supplementary Table S.2). Uptake was highest among men and women not living alone who were either retired or living with disability, across all education levels. For example, middle-educated women not living alone and with disability were 5.0 percentage points (ppts) above the mean (95% CI: 2.3 to 7.7). In contrast, uptake was lowest among lower- and middle-educated men, employed or unemployed, and among lower-educated retired men living alone. For example, lower-educated unemployed men living alone were 5.7 ppts (95% CI: -9.0 to - 2.3) below the mean.

**Table 3.**
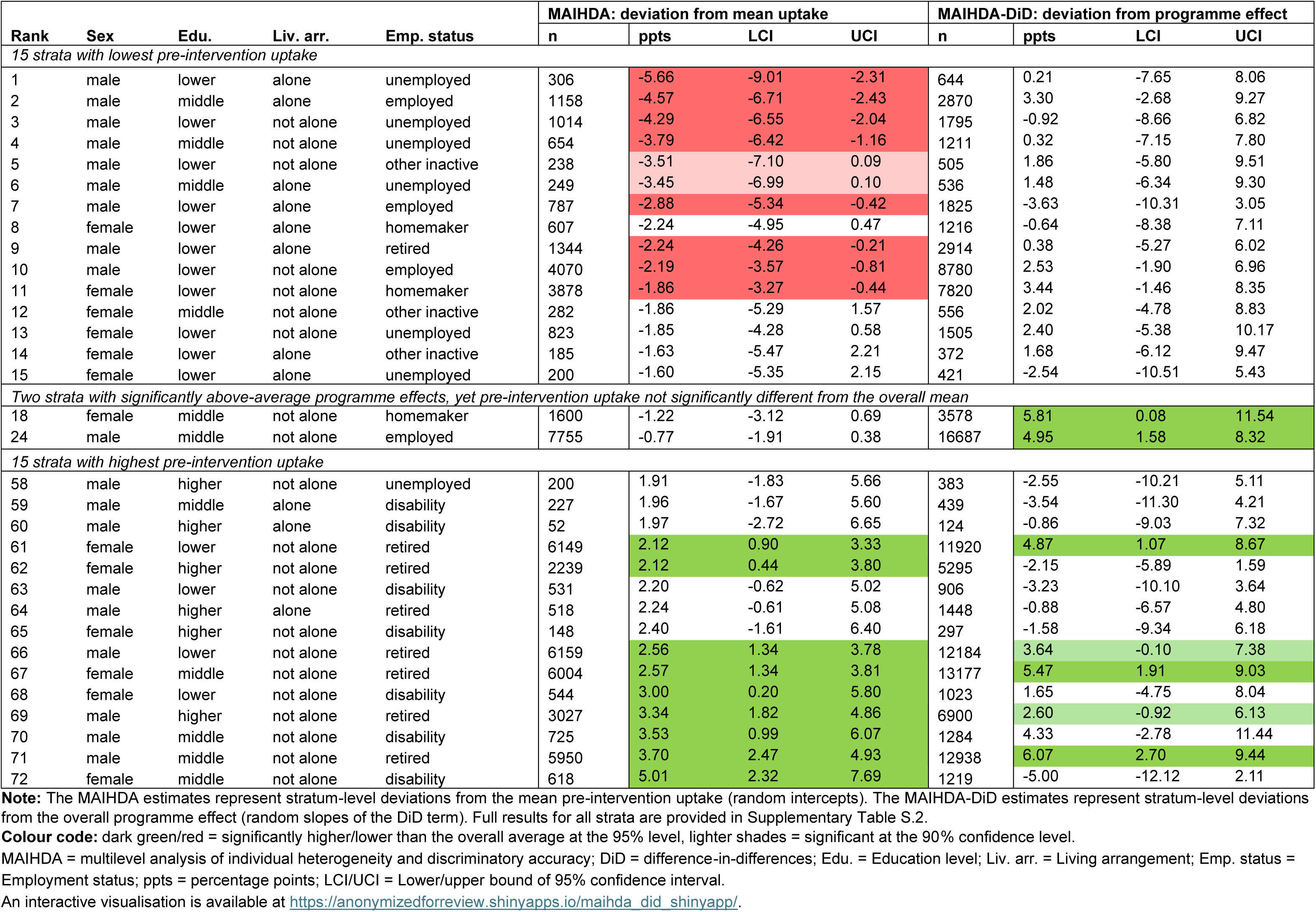
Predicted stratum-level deviation in CRC screening uptake from (1) the overall mean pre-intervention uptake (MAIHDA, EHIS-2) and (2) the overall programme effect (MAIHDA–DiD, EHIS-2 & EHIS-3), with 95% confidence intervals.

Although these patterns broadly aligned with the main effects, several strata deviated from these trends. For example, while men generally had lower uptake, retired men not living alone were among the highest-uptake strata. Homemaker women not living alone were the only female stratum with significantly below-average uptake. Similarly, although retirement and disability strata tended to have higher uptake, lower-educated retired men living alone screened less.

### Impact of CRC programmes across strata

Results from the MAIHDA-DiD regression are reported in Supplementary Table S.3. In Model 2a, the overall DiD coefficient indicated that CRC programme implementation increased past-two-year screening uptake by 25.6 ppts on average (95% CI: 15.9 to 35.2). Predicted programme effects across strata (random slopes of the DiD term) are shown in Table 3 (full results in Supplementary Table S.2). In addition, an interactive visualisation combining MAIHDA and MAIHDA-DiD stratum-level deviations is available at https://anonymizedforreview.shinyapps.io/maihda_did_shinyapp/.

Programme effects varied across strata. Several groups with higher pre-intervention uptake experienced above-average increases, including middle-educated men and women not living alone and retired, as well as lower-educated women in the same category. Two strata (middle-educated homemaker women and middle-educated employed men not living alone) also showed above-average increases although their pre-intervention uptake did not differ from the overall mean. Among strata with lower pre-intervention uptake, lower-educated employed men living alone experienced programme effects 3.6 ppts below the average effect; however, this deviation was not statistically significant, and their absolute screening uptake still increased substantially.

Models 2b-2e (Supplementary Table S.3) include interactions between the DiD term and each sociodemographic variable. PCV values indicate the extent to which each factor explained variation in programme effects across strata. Living arrangement accounted for more variation in the DiD random slope than other factors (PCV = 64.6%), with programme effects 6.2 ppts lower among individuals living alone than among those not living alone. Education explained 37.7% of the variance, with programme effects 5 ppts higher among middle-educated than higher-educated individuals. Employment status explained 31.6% of the variance, with effects 11.1 and 12.2 ppts lower for the unemployed and those with disability, compared with retirees. Sex had the lowest PCV (3.7%), and its interaction with the DiD term was not significant.

A fully saturated model including all main effects and interactions between the DiD term and the stratifying variables showed no evidence of multiplicative heterogeneity in programme effects, indicating that observed differences were explained by additive components (Model 2f, Supplementary Table S.3).

## Discussion

This study examined inequalities in CRC screening uptake across sociodemographic groups and assessed how population-based screening programmes affected these inequalities. It is the first study to combine MAIHDA with difference-in-differences methods (MAIHDA-DiD) to assess heterogeneous policy effects across groups defined by intersecting social conditions.

In the pre-intervention period, we observed inequalities consistent with well-established barriers to cancer screening participation, such as lower education, lower socioeconomic position and living alone, which likely reflect differences in health literacy, access to resources and social support (9, 11, 27). Gendered patterns were also evident, as men generally participate less in preventive healthcare and have poorer health-seeking behaviours (28).

The MAIHDA results shed light on subgroup-specific patterns shaped by intersecting social conditions that would be difficult to identify using conventional regression models treating sociodemographic factors as independent predictors. Screening uptake was highest among men and women not living alone, retired or living with disability, across all education levels. In contrast, the lowest uptake was observed among lower- and middle-educated men, whether unemployed or employed, and among lower-educated retired men living alone. These findings extend previous research documenting lower screening uptake among lower-educated groups (8), by showing that these inequalities were particularly pronounced among men. While a previous study found that men living alone were particularly disadvantaged in CRC screening (11), we also extend this evidence by identifying lower-educated men living alone as the most disadvantaged group. Additionally, lower-educated homemaker women were the only female subgroup with significantly below-average uptake, consistent with research linking gender inequality and domestic roles to lower screening uptake (29, 30).

While previous studies have reported mixed evidence on the association between employment status and CRC screening (10), its role became clearer when examined in combination with other social dimensions. Our results showed that retirement and disability coincided with above-average uptake primarily among individuals not living alone, whereas both employment and unemployment coincided with below-average uptake among lower-educated men. These findings illustrate how intersecting conditions jointly shape social positions, producing cumulative (dis)advantages in preventive healthcare use.

A key contribution of this study is the assessment of CRC programme effects on screening inequalities using MAIHDA-DiD. Exploiting cross-country policy variation, we found that these programmes substantially increased overall screening uptake, consistent with prior research (21). However, programmes did not produce above-average increases among groups with lower pre-intervention uptake and therefore did not reduce inequalities. Instead, they generated above-average increases among groups with already higher uptake, such as middle-educated men and women not living alone and retired, and lower-educated women in the same category, thereby widening inequalities. Importantly, although middle-educated groups benefited more from the programmes than higher-educated groups, the largest gains were observed among middle-educated groups that had higher pre-intervention uptake, namely those not living alone and retired. In contrast, middle-educated groups with lower pre-intervention uptake, such as unemployed men living alone, benefited comparatively less.

These findings suggest that some groups are better positioned to respond to programme invitations, likely due to greater social support and fewer structural barriers, such as time constraints or competing demands. Consequently, population-based programmes may unintentionally amplify existing social gradients, improving overall uptake without reducing (and sometimes widening) inequalities, a pattern observed across public health interventions more broadly (31, 32). From a vulnerable populations perspective, social structures shape subgroups’ social position and risk profiles, yet interventions often fail to address the underlying unequal access to resources (e.g., knowledge, social support, discretionary time) that enable individuals to benefit from population-level interventions (33). While CRC programmes have increased screening coverage across the eligible population, they are unlikely to narrow existing gaps without explicit equity-oriented components. Evidence from systematic reviews suggests that combining mailed outreach with strategies such as reminders, general practitioner endorsement or patient navigation can improve uptake among underserved populations (34-36). As Europe’s Beating Cancer Plan targets 90% screening participation (5), monitoring subgroup inequalities remains essential to ensure equitable progress.

Our study has several limitations. We focused on four sociodemographic dimensions and alternative classifications may yield different insights. Other relevant factors, such as ethnicity, migration status or household structure were unavailable or inconsistently measured across EHIS waves. Future research should consider incorporating ethnicity, given its relevance for CRC outcomes (37). We could have considered additional dimensions, such as nationality or marital status; however, incorporating more categories would substantially increase the number of strata, many of which would contain very few or no individuals, making results difficult to present and interpret. Screening uptake was self-reported and may be affected by recall and social desirability bias, although prior studies suggest reasonable accuracy (38, 39). As EHIS did not collect information on respondents’ cancer history or reasons for screening, we could not distinguish preventive screening from diagnostic or symptom-related screening, potentially inflating our measure of preventive uptake. However, this is unlikely to bias DiD estimates, which rely on changes over time rather than absolute screening levels. As the pre-post intervention timeframe was relatively short, future studies should assess the longer-term effects of programmes on screening inequalities. Finally, although programme implementation was observed in only four countries, the findings are likely relevant to other European and high-income countries considering mailed FIT programmes, as similar social structures and screening delivery models underpin inequalities in screening uptake across contexts.

A key strength of this study is the innovative combination of MAIHDA with difference-in-differences methods, enabling the assessment of heterogeneous programme effects while accounting for the multilevel and sociodemographic structure of the data. Whereas conventional single-level regressions require several interaction terms to capture subgroup heterogeneity, MAIHDA summarises between-stratum variation with a single variance component, offering a more parsimonious and scalable approach to modelling complex patterns of inequality (13). Moreover, an advantage of multilevel models is shrinkage (partial pooling), whereby information is shared across strata, improving estimation for small subgroups and reducing the risk of spurious results (40). This results in more stable and reliable estimates than conventional regression models with interaction terms. As such, the approach could be used to consider heterogeneity in treatment effects, in social epidemiology, sociology of health and other disciplines.

## Conclusions

By combining MAIHDA with difference-in-differences methods (MAIHDA-DiD), this study introduces a novel framework for assessing heterogeneous effects of policy interventions across population groups. Applied to CRC screening programmes introduced in four European countries, MAIHDA-DiD showed that although programmes substantially increased screening uptake overall, they did not produce above-average gains among groups with lower pre-intervention uptake and therefore did not reduce inequalities. Instead, inequalities widened, with larger increases observed among groups that already had higher uptake. Living arrangement was a key determinant of unequal programme benefits, with individuals living alone benefiting less. These findings highlight the need to complement population-based screening programmes with targeted, equity-oriented strategies to ensure that advances in cancer prevention benefit all social groups.

## Author contributions

Conceptualisation: V.J.; Data curation: V.J.; Formal analysis: V.J.; Interpretation of data: V.J., K.D., I.K., S.C., A.B., D.H.; Funding acquisition: V.J., S.C., A.B.; Investigation: V.J.; Methodology: V.J., A.B.; Project administration: V.J., S.C.; Supervision: V.J., I.K., S.C., D.H.; Validation: V.J., S.C.; Visualisation: V.J.; Writing - original draft preparation: V.J.; Writing - review & editing: V.J., K.D., I.K., S.C., A.B., D.H.; All authors have read and approved the submitted version.

## Ethics approval

This observational study used anonymised data provided by Eurostat and did not require ethical approval.

## Declaration of generative AI and AI-assisted technologies in the writing process

During the preparation of this manuscript, generative AI (ChatGPT, GPT-5.2, OpenAI, December 2025 version) was used only to improve language and clarity under full human oversight and control. No AI tools were used to generate scientific content, analyse or interpret data, or draw conclusions. All content was reviewed and edited by the authors.

## Data availability statement

Data from the European Health Interview Survey are publicly available at no cost; however, access to anonymised microdata requires a formal application to Eurostat. Information on how to request access can be found at https://ec.europa.eu/eurostat/web/microdata/european-health-interview-survey.

## Acknowledgements

We acknowledge the support of Eurostat in granting access to the anonymised EHIS data used in this study. We would like to thank the following experts, researchers, and stakeholders across Europe for providing information on the characteristics of national colorectal cancer screening programmes: Michel Candeur, Inge Wauters (Belgium); Sisse Helle Njor, Linn Freund, Tina Bech Olesen, Henry Jensen, Holger Schildt Knudsen, Kristoffer Lande Andersen (Denmark); Elle Langens, Femke Sijtsma (Netherlands); Claire Dillenbourg, Fanny Lorin (Luxembourg); Therese Mooney (Ireland); Renata Chloupkova on behalf of the National Screening Centre (Czech Republic); Sascha Reiff (Malta); Eliise Leif (Estonia); and Marcell Csanadi (Hungary). The information they shared was essential for understanding and accurately classifying the colorectal cancer screening programmes of the countries included in this study.

