## Supplementary Appendix A for "Inequalities in Colorectal Cancer Screening: Combining MAIHDA with Difference-in-Differences to Assess Programme Effects Across Population Subgroups"

### ONLINE APPENDIX

#### Appendix A: Difference-in-differences (DiD) model assumptions

*Parallel trends assumption*

The DiD model compares changes in screening uptake between countries that introduced screening programmes with mailed FIT kits (intervention group) and those that did not (comparison group). For causal identification, the DiD approach relies on the parallel trends assumption, namely that, in the absence of programme implementation, differences in screening uptake between intervention and comparison groups would have remained constant over time. Under this framework, common temporal trends in screening uptake are accounted for, and the model controls for both observed and unobserved time-invariant differences between intervention and comparison countries. If screening uptake followed parallel trends prior to programme implementation, and if implementation was not driven by pre-existing trends, any divergence observed after implementation can be interpreted as the effect of the screening programmes.

Pre-intervention trends could be assessed for only one intervention country (Belgium) and 10 comparison countries, as fewer countries participated in the earlier EHIS-1 wave, and Austria and Estonia did not collect information on stool-based screening uptake in EHIS-1. As shown in Figure A.1, trends in past-two-year screening uptake were parallel between Belgium and the comparison group during the pre-intervention period. Following programme implementation, screening uptake increased markedly in Belgium, diverging from the comparison group, as expected. Additionally, we performed a regression-based test of pre-intervention trends (Table A.1). Screening uptake was regressed on survey wave, the intervention variable (Belgium versus comparison group) and their interaction. The interaction term between Belgium and EHIS-2 was small and not significant ($b = 0.008$, $p = 0.928$), indicating no differential trends between Belgium and the comparison group prior to programme implementation and thereby supporting the parallel trends assumption.

Finally, pre-intervention screening uptake levels in EHIS-2 were very similar between intervention and comparison groups (approximately 22%; Table A.2), based on the full analytical sample of 24 countries, providing further support for the plausibility of the parallel trends assumption.

**Figure A.1 Pre-intervention trends in stool-based screening uptake within the past two years**

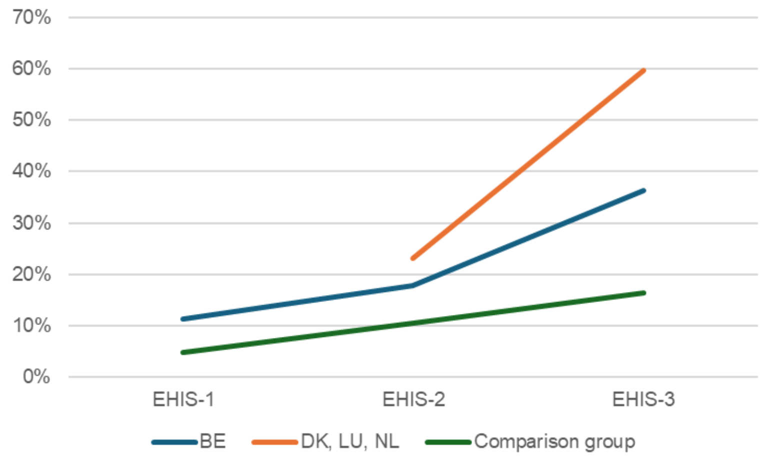


Note: The comparison group included 10 countries that took part in all three EHIS waves and for which data on stool-based screening uptake was available. Denmark, Luxembourg and The Netherlands did not participate in EHIS-1.
EHIS = European Health Interview Survey; BE = Belgium; DK = Denmark; LU = Luxembourg; NL = The Netherlands.

**Table A.1 Test of pre-intervention trends in stool-based screening uptake, Belgium (intervention) versus comparison group of countries**

| | b | 95% CI | p-value |
|---|---|---|---|
| Intercept | 0.067 | 0.013, 0.121 | 0.015 |
| Belgium (ref: comparison group) | 0.045 | -0.134, 0.224 | 0.624 |
| Survey wave (ref.: EHIS-1) | | | |
| EHIS-2 | 0.058 | 0.007, 0.108 | 0.025 |
| EHIS-3 | 0.105 | 0.055, 0.155 | <0.001 |
| Belgium x survey wave (ref.: EHIS-1) | | | |
| Belgium x EHIS-2 | 0.008 | -0.159, 0.175 | 0.928 |
| Belgium x EHIS-3 | 0.146 | -0.021, 0.313 | 0.086 |
| *Random intercept variances* | | | |
| Country-wave level | 0.003 | | |
| Country level | 0.004 | | |
| Individual level residual variance | 0.089 | | |
| N | 126,744 | | |

Note: the comparison group included 10 countries that took part in all three EHIS waves and for which data on stool-based screening uptake was available.
CI = confidence interval.

**Table A.2 Stool-based screening uptake within the past two years, before and after CRC programme implementation, EHIS-2 and EHIS-3 (N = 201,214)**

| | Pre-intervention (EHIS-2) | Post-intervention (EHIS-3) | Overall |
|---|---|---|---|
| | % (n) | % (n) | % (n) |
| Comparison group (no programme) | 21.9 (18,859) | 27.5 (27,112) | 24.9 (45,971) |
| Intervention group (programme) | 22.0 (1,642) | 55.6 (5,107) | 40.6 (6,749) |
| Overall | 21.9 (20,501) | 29.9 (32,219) | 26.2 (52,720) |

EHIS = European Health Interview Survey

*Falsification and placebo tests*

To further assess the validity of the difference-in-differences (DiD) identification strategy, we conducted falsification and placebo tests using data from EHIS-2 and EHIS-3. Falsification tests applied the DiD analysis to populations not eligible for CRC programmes, namely individuals aged 30-49 and 80 years and older (Table A.3). No programme effects were found in these groups, indicating that the intervention did not influence screening uptake outside the target population (50-74 years old). This finding supports the interpretation that the observed effects among the eligible individuals are attributable to the intervention itself rather than to broader underlying trends. In addition, we conducted placebo tests by replicating the DiD models using outcomes that should not be affected by CRC programmes, specifically cholesterol and blood sugar checks (Table A.4). As expected, no effects were detected for these outcomes. Overall, these tests provided strong support for the validity of the DiD identification strategy, reinforcing the interpretation that the estimated programme effects reflect the impact of CRC programmes rather than unmeasured confounding, secular trends or broader changes in healthcare utilisation.

**Table A.3 Effect of CRC programmes on stool-based screening uptake within the past two years, among older (80+) and younger (30-49) age groups**

| | b | 95% CI | p-value |
|---|---|---|---|
| **80 years old and older** | | | |
| Intercept | 0.154 | 0.106, 0.202 | <0.001 |
| Programme (ref.: no programme) | -0.022 | -0.137, 0.092 | 0.704 |
| Post-intervention (ref.: pre-intervention period) | 0.013 | -0.008, 0.034 | 0.225 |
| DiD | 0.005 | -0.048, 0.058 | 0.843 |
| *Random intercept variances* | | | |
| Country-wave level | 0.001 | | |
| Country level | 0.010 | | |
| Individual level residual variance | 0.125 | | |
| N | 27,684 | | |
| **30-49 years old** | | | |
| Intercept | 0.074 | 0.047, 0.101 | <0.001 |
| Programme (ref.: no programme) | 0.013 | -0.054, 0.080 | 0.706 |
| Post-intervention (ref.: pre-intervention period) | 0.014 | 0.003, 0.025 | 0.010 |
| DiD | -0.018 | -0.045, 0.009 | 0.193 |
| *Random intercept variances* | | | |
| Country-wave level | 0.000 | | |
| Country level | 0.004 | | |
| Individual level residual variance | 0.077 | | |
| N | 148,061 | | |

Notes: DiD (difference-in-differences) denotes the *Programme x Post-intervention* interaction term.
CI = confidence interval.

**Table A.4 Effect of CRC programmes on placebo outcomes**

| | b | 95% CI | p-value |
|---|---|---|---|
| **Cholesterol check (past year uptake)** | | | |
| Intercept | 0.643 | 0.593, 0.693 | <0.001 |
| Programme (ref.: no programme) | 0.022 | 0.004, 0.039 | 0.014 |
| Post-intervention (ref.: pre-intervention period) | 0.024 | -0.099, 0.146 | 0.706 |
| DiD | -0.023 | -0.067, 0.020 | 0.298 |
| *Random intercept variances* | | | |
| Country-wave level | 0.001 | | |
| Country level | 0.012 | | |
| Individual level residual variance | 0.212 | | |
| N | 199,674 | | |
| **Blood sugar check (past year uptake)** | | | |
| Intercept | 0.644 | 0.597, 0.691 | <0.001 |
| Programme (ref.: no programme) | 0.010 | -0.105, 0.125 | 0.867 |
| Post-intervention (ref.: pre-intervention period) | 0.022 | 0.003, 0.040 | 0.020 |
| DiD | -0.021 | -0.067, 0.024 | 0.361 |
| *Random intercept variances* | | | |
| Country-wave level | 0.001 | | |
| Country level | 0.011 | | |
| Individual level residual variance | 0.214 | | |
| N | 198,982 | | |

Notes: DiD (difference-in-differences) denotes the *Programme x Post-intervention* interaction term.
CI = confidence interval.
