## Supplementary Tables for "Inequalities in Colorectal Cancer Screening: Combining MAIHDA with Difference-in-Differences to Assess Programme Effects Across Population Subgroups"

### ONLINE SUPPLEMENTARY MATERIALS

**Table S.1 Classification of countries into intervention and comparison groups for the MAIHDA-DiD model**

| | Comparison | Intervention |
|---|---|---|
| AT | X | |
| BE | | X |
| BG | X | |
| CY | X | |
| DE | X | |
| DK | | X |
| EE | X | |
| EL | X | |
| ES | X | |
| FI | X | |
| HU | X | |
| IS | X | |
| IT | X | |
| LT | X | |
| LU | | X |
| LV | X | |
| MT | X | |
| NL | | X |
| NO | X | |
| PL | X | |
| PT | X | |
| RO | X | |
| SE | X | |
| SK | X | |

Note: Intervention countries implemented nationwide CRC screening programmes with mailed FIT kits for all eligible individuals during the period between the second (EHIS-2) and the third (EHIS-3) waves of the European Health Interview Survey. Although Luxembourg initially launched a pilot programme, it was classified as a full programme because FIT kits were mailed to all eligible individuals between 2014 and 2019 (Rossini, 2018).
Countries were selected based on data availability before and after the intervention period, allowing a quasi-experimental design. The United Kingdom did not participate in EHIS-3, and although France participated, its data was not released by Eurostat. Slovenia, Ireland and Croatia were excluded because their mailed-kit programmes predated the intervention period. Czechia was excluded because its programme relied on mailed invitation letters without FIT kits.
MAIHDA = Multilevel Analysis of Individual Heterogeneity and Discriminatory Accuracy; DiD = Difference-in-Differences

**Table S.3 MAIHDA-DiD regression estimates for past-two-year CRC screening uptake, EHIS-2 and EHIS-3 (N= 201,214)**

| | Model 2a | | Model 2b | | Model 2c | | Model 2d | | Model 2e | | Model 2f* | |
|---|---|---|---|---|---|---|---|---|---|---|---|---|
| | Estimate | 95% CI | Estimate | 95% CI | Estimate | 95% CI | Estimate | 95% CI | Estimate | 95% CI | Estimate | 95% CI |
| **Fixed-effects** | | | | | | | | | | | | |
| Intercept | 0.1730 | 0.1100, 0.2360 | 0.1791 | 0.1156, 0.2425 | 0.1924 | 0.1288, 0.2560 | 0.1814 | 0.1181, 0.2447 | 0.1882 | 0.1239, 0.2524 | 0.2203 | 0.1568, 0.2837 |
| Programme (ref.: no programme) | | | | | | | | | | | | |
| programme | 0.0341 | -0.1191, 0.1873 | 0.0235 | -0.1300, 0.1769 | 0.0255 | -0.1280, 0.1791 | 0.0348 | -0.1184, 0.1880 | 0.0320 | -0.1219, 0.1859 | 0.0081 | -0.1469, 0.1630 |
| Time (ref.: pre-intervention) | | | | | | | | | | | | |
| post-intervention | 0.0444 | 0.0053, 0.0835 | 0.0466 | 0.0072, 0.0861 | 0.0472 | 0.0078, 0.0867 | 0.0452 | 0.0059, 0.0845 | 0.0540 | 0.0138, 0.0942 | 0.0584 | 0.0184, 0.0984 |
| DiD (Programme x Time) | 0.2559 | 0.1593, 0.3524 | 0.2628 | 0.1648, 0.3609 | 0.2796 | 0.1812, 0.3780 | 0.2831 | 0.1867, 0.3794 | 0.2265 | 0.1265, 0.3265 | 0.2814 | 0.1817, 0.3812 |
| **DiD x sociodemographic variables** | | | | | | | | | | | | |
| *Sex (ref.: female)* | | | | | | | | | | | | |
| male | | | -0.0131 | -0.0299, 0.0038 | | | | | | | -0.0127 | -0.0210, -0.0044 |
| Time x male | | | -0.0049 | -0.0170, 0.0071 | | | | | | | -0.0073 | -0.0151, 0.0004 |
| Programme x male | | | 0.0219 | 0.0027, 0.0411 | | | | | | | 0.0257 | 0.0058, 0.0457 |
| DiD x male | | | -0.0130 | -0.0514, 0.0253 | | | | | | | -0.0062 | -0.0329, 0.0205 |
| *Education (ref.: higher)* | | | | | | | | | | | | |
| low | | | | | -0.0295 | -0.0498, -0.0092 | | | | | -0.0248 | -0.0359, -0.0137 |
| middle | | | | | -0.0134 | -0.0336, 0.0069 | | | | | -0.0088 | -0.0195, 0.0019 |
| Time x low | | | | | -0.0184 | -0.0330, -0.0039 | | | | | -0.0162 | -0.0275, -0.0049 |
| Time x middle | | | | | -0.0064 | -0.0204, 0.0075 | | | | | -0.0053 | -0.0154, 0.0048 |
| Programme x low | | | | | 0.0086 | -0.0164, 0.0335 | | | | | 0.0106 | -0.0151, 0.0363 |
| Programme x middle | | | | | -0.0019 | -0.0253, 0.0216 | | | | | -0.0011 | -0.0246, 0.0225 |
| DiD x low | | | | | 0.0418 | -0.0035, 0.0870 | | | | | 0.0344 | -0.0004, 0.0692 |
| DiD x middle | | | | | 0.0503 | 0.0066, 0.0939 | | | | | 0.0476 | 0.0164, 0.0788 |
| *Living arrangement (ref.: not alone)* | | | | | | | | | | | | |
| alone | | | | | | | -0.0187 | -0.0354, -0.0020 | | | -0.0204 | -0.0292, -0.0116 |
| Time x alone | | | | | | | -0.0014 | -0.0142, 0.0114 | | | 0.0001 | -0.0091, 0.0093 |
| Programme x alone | | | | | | | -0.0029 | -0.0263, 0.0206 | | | 0.0000 | -0.0238, 0.0238 |
| DiD x alone | | | | | | | -0.0617 | -0.0977, -0.0256 | | | -0.0611 | -0.0930, -0.0291 |
| *Employment status (ref.: retired)* | | | | | | | | | | | | |
| employed | | | | | | | | | -0.0302 | -0.0475, -0.0129 | -0.0317 | -0.0415, -0.0219 |
| unemployed | | | | | | | | | -0.0455 | -0.0667, -0.0244 | -0.0508 | -0.0659, -0.0357 |
| disability | | | | | | | | | 0.0167 | -0.0055, 0.0389 | 0.0184 | 0.0019, 0.0348 |
| homemaker | | | | | | | | | -0.0306 | -0.0546, -0.0066 | -0.0389 | -0.055, -0.0227 |
| other inactive | | | | | | | | | -0.0399 | -0.0668, -0.0130 | -0.0427 | -0.0648, -0.0206 |
| Time x employed | | | | | | | | | 0.0008 | -0.0111, 0.0127 | 0.0014 | -0.0071, 0.0098 |
| Time x unemployed | | | | | | | | | -0.0174 | -0.0377, 0.0029 | -0.0167 | -0.035, 0.0015 |

| | | | | | | | | |
|---|---|---|---|---|---|---|---|---|
| Time x disability | | | | | 0.0073 | -0.0154, 0.0299 | 0.0087 | -0.0122, 0.0296 |
| Time x homemaker | | | | | -0.0276 | -0.0475, -0.0078 | -0.0279 | -0.044, -0.0119 |
| Time x other inactive | | | | | 0.0192 | -0.0102, 0.0486 | 0.0198 | -0.0081, 0.0477 |
| Programme x employed | | | | | 0.0129 | -0.0088, 0.0345 | 0.0152 | -0.0068, 0.0371 |
| Programme x unemployed | | | | | 0.0645 | 0.0093, 0.1197 | 0.0682 | 0.0131, 0.1234 |
| Programme x disability | | | | | 0.0184 | -0.0315, 0.0684 | 0.0185 | -0.0316, 0.0685 |
| Programme x homemaker | | | | | 0.0091 | -0.0310, 0.0492 | 0.0202 | -0.0219, 0.0623 |
| Programme x other inactive | | | | | 0.0254 | -0.0271, 0.0778 | 0.0305 | -0.022, 0.0831 |
| DiD x employed | | | | | -0.0224 | -0.0639, 0.0190 | -0.0264 | -0.0555, 0.0028 |
| DiD x unemployed | | | | | -0.1105 | -0.1942, -0.0268 | -0.1160 | -0.1946, -0.0373 |
| DiD x disability | | | | | -0.1220 | -0.1964, -0.0476 | -0.1266 | -0.1948, -0.0584 |
| DiD x homemaker | | | | | 0.0374 | -0.0338, 0.1085 | 0.0104 | -0.0502, 0.0709 |
| DiD x other inactive | | | | | -0.0260 | -0.1049, 0.0528 | -0.0387 | -0.1114, 0.034 |
| **Random intercepts variances** | | | | | | | | |
| Country-time level | 0.0039 | 0.0039 | 0.0038 | 0.0039 | 0.0039 | | 0.0038 | |
| Country level | 0.0164 | 0.0164 | 0.0164 | 0.0164 | 0.0164 | | 0.0164 | |
| Stratum level | 0.0009 | 0.0009 | 0.0004 | 0.0008 | 0.0008 | | 0.0000 | |
| **Random slopes variances (stratum-level)** | | | | | | | | |
| DiD | 0.0019 | 0.0018 | 0.0012 | 0.0007 | 0.0013 | | 0.0000 | |
| Time | 0.0002 | 0.0002 | 0.0001 | 0.0002 | 0.0001 | | - | |
| Individual level residual variance | 0.1625 | 0.1625 | 0.1625 | 0.1625 | 0.1625 | | 0.1625 | |
| **PCV of DiD random slope (%)** | - | 3.7% | 37.7% | 64.6% | 31.6% | | 100.0% | |

Note: Multilevel model with individuals cross-classified by sociodemographic strata (n = 72), countries (n = 24) and country-time clusters (n = 48).
MAIHDA = multilevel analysis of individual heterogeneity and discriminatory accuracy; DiD = difference-in-differences; PCV = proportional change in variance
*Model 2f did not converge when allowing for random slopes for time and the DiD term at the stratum level simultaneously. To achieve model convergence, we removed the random slope for time while retaining the DiD random slope.
